# Health Services Interventions to Improve the Quality of Care in Rare Disease: A Scoping Review

**DOI:** 10.1101/2024.02.07.24302315

**Authors:** Cody Chou, Sydney O. Wiredu, Liesel Von Imhof, Anran Tan, Sasha Agarwal, Melis Lydston, Vanessa L. Merker

**Affiliations:** Harvard College, Cambridge, MA, 02138; Massachusetts General Hospital, Boston, MA, 02144; Harvard School of Public Health, Boston, MA, 02115; Harvard Medical School, Boston, MA, 02115

**Keywords:** scoping review, rare diseases, rare disorders, implementation science, health services research

## Abstract

**Background:** Rare diseases are often complex, multi-system disorders requiring specialized, lifelong care. These diseases share significant challenges in healthcare delivery, including diagnostic delays, limited access to specialists, and lack of effective treatments.

**Objectives:** To aggregate and critically examine innovative health services interventions for rare diseases, with the goal of identifying effective, scalable strategies to improve care

**Design:** Scoping review

**Data Sources:** Comprehensive searches were conducted in April 2022 in Ovid MEDLINE, Embase.com, Web of Science Core Collection, Cochrane CENTRAL, and ClinicalTrials.gov.

**Methods:** We sought to identify peer-reviewed original research published in English reporting results of interventions to improve guideline-concordant care, care coordination, and care transitions in rare disease populations. Using Covidence software, two researchers independently screened abstracts using pre-established inclusion and exclusion criteria, with conflicts resolved through consensus discussion with a third researcher. The same process was used to screen full-text research for eligibility and to extract study characteristics and results from eligible research.

**Results:** Our search identified 2899 articles. After screening for eligibility criteria, 12 articles describing health services interventions in rare diseases were identified. Most studies were conducted in Europe and involved adult participants. Three main intervention targets were identified: 1) increasing patients’ access to multidisciplinary expertise (e.g. using expert panels/tumor boards, integrating additional disciplines into care teams, and creating a hotline for specialist advice); 2) using technology to give point-of-care physicians access to information (e.g., electronic medical record templates/decision-support), and 3) standardizing care through clinical care pathways.

**Conclusions:** This review identified several efficacious interventions to improve healthcare delivery for individuals with a single rare disease. Testing these strategies across broader groups of rare disease patients could more efficiently improve healthcare delivery for the rare disease community, facilitating patients’ timely access to treatments, improving their health outcomes, and potentially reducing healthcare costs with economies of scale.

**Plain Language Summary:** *Why was this study done?:* While each rare disease is unique and affects only a small number of individuals, the rare disease community as a whole faces similar challenges seeking healthcare, such as delayed diagnosis, limited access to specialists, and insufficient treatment options. This study seeks to find innovative strategies to enhance healthcare delivery that have been tested in at least one rare disease that might be applicable across a broad spectrum of these conditions.

*What did the researchers do?:* We reviewed public reports of research that aimed to improve guideline-concordant care, care coordination, or care transitions for people with rare diseases. At least two researchers screened each paper to assess whether it met inclusion and exclusion criteria, and all conflicts were resolved by consensus discussion.

*What did we find?:* After searching 2899, we found 12 describing successful interventions for people with rare diseases. These interventions either 1) increased patients’ access to multidisciplinary expertise (through things like expert review panels, integrating pharmacists into the care team, or creating a specialist hotline); 2) used technology to facilitate physicians access to rare disease information (through things like electronic medical record templates and alerts); or 3) standardize care delivery through creating set clinical pathways.

*What do these findings mean?:* These results highlight how efforts to improve access to multidisciplinary experts, implement new technology, and standardize care for rare disease could be used to develop impactful healthcare interventions that are transferable across multiple rare diseases.

## Introduction

Rare diseases, which are defined in the U.S. as diseases affecting less than 200,000 Americans and in the European Union as diseases affecting no more than 1 out of 2,000 people, are often complex, multi-system disorders that require lifelong, specialized healthcare and support.^1, 2^ There are between 5,000 and 8,000 distinct rare diseases, affecting 6-8% of the population in total.^3^ Taken together, these diseases cause significant morbidity and mortality, negatively impact quality of life, and can confer a tremendous social and economic burden on families and communities.^4^

In recent years, there have been many therapeutic advances to improve the health of patients living with rare conditions. Due to the demographic and symptomatic heterogeneity of rare diseases, these advances have often only been studied in individual diseases or closely related groups of diseases.^5^ But rare diseases also share many common health challenges, such as delays in diagnosis, limited access to specialists, and experience of societal stigma.^6^ Developing healthcare delivery interventions that could address these common challenges simultaneously across multiple rare diseases could amplify the impact of rare disease research, optimizing the often limited resources within rare disease communities while also fostering a more inclusive and comprehensive approach to rare disease care.

However, the body of literature on rare diseases is often dispersed across specialty- or disease-specific journals, making it difficult to generalize novel health services intervention methods across a range of rare diseases to improve patient health outcomes. For this reason, we undertook a scoping review to systematically search for and analyze healthcare delivery interventions used to improve care for rare diseases, with the goal of revealing promising strategies that could potentially be adapted to address a wider spectrum of rare diseases. We focused our review on three common challenges to providing care for rare diseases: care coordination, care transitions, and receiving guideline-concordant care (or when formal guidelines are lacking, care according to expert-recommended best practices).

Care coordination is especially challenging for rare disease patients due to their need for specialized care across multiple providers and settings.^7, 8^ Individuals with rare diseases and their family members report often having to relay information between various health professionals, attend numerous health appointments at different locations, and encounter inadequate communication and health data sharing among healthcare providers.^9^ Furthermore, as more children and adolescents survive into adulthood, a related challenge arises involving transitioning from pediatric to adult clinics, and coordinating the exchange of information between these providers.^10, 11^ As rare disease patients transition from parent-supervised pediatric care to more independent adult models, they may face challenges taking over their complex care. For example, some patients struggle with the work involved in transferring medical records and a lack of trust in adult providers.^12^ Transitions of care between inpatient and outpatient settings may also be required as the disease progresses between stages or requires the involvement of different specialists.

Finally, many patients with rare disease have difficulty accessing specialists with adequate knowledge of their rare disease.^13^ While there is extensive literature on best practices and clinical guidelines for common diseases, recommendations targeted at rare diseases remain relatively sparse and may be hard for front-line clinicians to access and act on amidst the pressures of increasingly short healthcare visits. However, interventions to facilitate access to information on clinical best practices could help local care teams treat these complex and unfamiliar issues specific to rare diseases.^14^ Based on the potential for cross-disorder health services interventions to remedy these common challenges in quality of care, we undertook the following scoping review to locate proven interventions for patients with rare diseases, analyze common intervention mechanisms, and discuss the potential for interventions to be translated to other rare diseases.

## Methods

After refining our objective above, we designed a scoping review protocol. Standard systematic reviews are typically designed to analyze a narrow range of studies, where relevant study designs can often be identified in advance.^15^ However, because we wanted to include a variety of populations, interventions, and study designs, a scoping review was deemed most appropriate. Scoping reviews also have the ability to identify possible gaps in the evidence base.^15^ We then refined our research question to focus on three major issues that affect access to and/or quality of treatment for rare disease: guideline-concordant/expert-recommended care, care coordination, and care transitions. From these common challenges, we aimed to systematically identify health services interventions innovating new processes of care that could apply across multiple diseases, and excluded medical, surgical, or psychological interventions focused narrowly on biologic outcomes specific to unique rare disease processes.

### Identifying relevant studies (literature search)

Electronic searches for published literature were conducted by a medical librarian in April 2022 using Ovid MEDLINE (1946 to present), Embase.com (1947 to present), Web of Science Core Collection (1900 to present), Cochrane Central Register of Controlled Trials (CENTRAL) via Ovid (1991 to present) and ClinicalTrials.gov (1999 to present). The search strategy was designed by an experienced research librarian (ML) and incorporated controlled vocabulary and free-text synonyms for the concepts of rare diseases, care coordination, care transitions, and guideline-concordant care. The full database search strategies are documented in Appendix A. The search strategy for coordinated care was adapted from a previous search by McDonald et al.^7^ No restrictions on language or any other search filters were applied. All identified studies were combined and de-duplicated in a single reference manager (EndNote).

The citations were then uploaded into Covidence review software for screening.^16^

### Study selection

Two researchers screened the titles and abstracts of retrieved records to determine if the papers potentially met the study’s inclusion and exclusion criteria. We included publications if they: 1) studied a disease that is rare within the studied country (i.e. affected less than 200,000 Americans if conducted in the U.S. or had a prevalence less than 1 in 2000 in all other countries); 2) studied a health services research intervention or policy that aimed to improve guideline-concordant/expert-recommended care, care transitions (including patient navigation) and/or care coordination (including integrated care); 3) was an original research article or a clinical trial with published results; and 4) was published in the English language. We excluded articles if they: 1) studied medical, surgical, psychological, or lifestyle interventions (rather than healthcare delivery interventions); 2) focused solely on creation of disease-specific guidelines, creation of patient registries, or diagnosis of individuals with rare diseases (rather than their ongoing care/treatment); 3) were individual case reports, conference abstracts, editorials, or commentaries. All conflicts between the two researchers were discussed as a group with the senior researcher and resolved by consensus. All potentially eligible manuscripts from this stage were retrieved and advanced to full-text screening, which followed an identical process of review against inclusion/exclusion criteria by two independent researchers with conflicts resolved during consensus discussion with the senior researcher.

### Data Extraction and Analysis

Two researchers independently extracted data from eligible full-texts using a standardized template within Covidence. Any conflicts were reviewed and adjudicated by a third researcher. Papers were descriptively summarized by extracting the following study characteristics: study location and timing, study aim, study design, participant demographics, description of interventions, study outcome measures and study results. After data was extracted, the research team reviewed included interventions to inductively create themes describing common intervention strategies.

## Results

The search strategy and study selection process are outlined in Figure 1. Of 2,899 records screened based on title and abstract, 80 full-text articles were assessed for eligibility and 12 articles were included in the final review. A majority of the studies included sites in Europe (n=8); studies also included sites in the United States (n=6), Asia (n=1), and Africa (n=1). Included studies assessed a wide variety of rare diseases, including sarcomas (2), blood disorders (4), lung conditions (2), autoimmune or inherited disorders (2), brain disorders (2), and cardiac disorders (1). Only one included article described a randomized controlled trial; the remaining studies included 7 prospective observational studies and 5 retrospective studies. A majority of studies included only adults (5) or both adults and children (4); two included only pediatric populations and one did not specify demographics of the study population. Seven studies focused on guideline-concordant care, six studies focused on care coordination, and two focused on care transitions (more than one focus was present in two studies). Additional details of the included studies are summarized in Table 1 and below.

**Figure 1.**
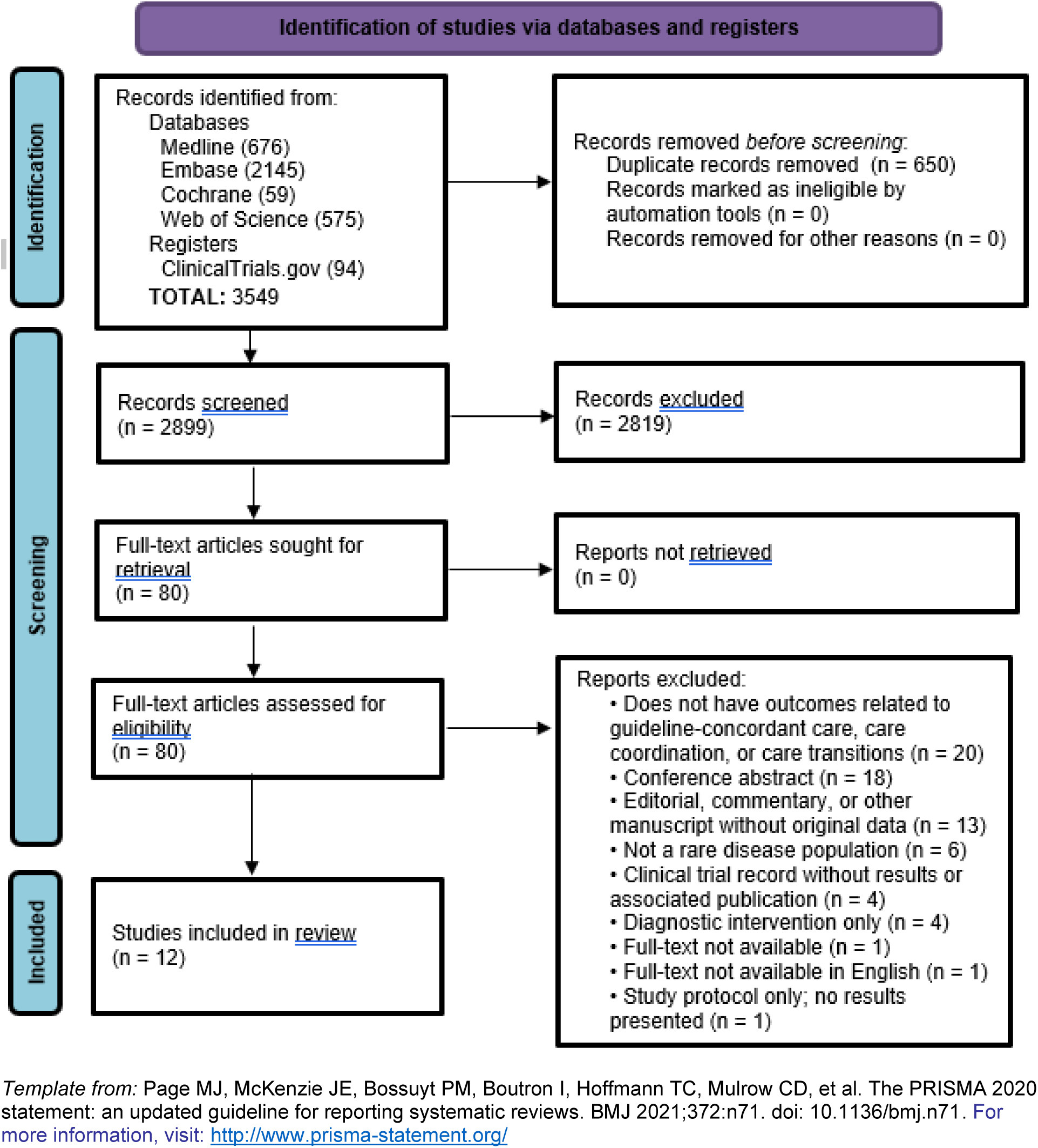
PRISMA Flow Diagram.

**Table 1.** Characteristics of Included Studies.

| Report | Rare Disease | Study Population<sup>1</sup> | Description of Intervention | Study Outcome Measures/Results<sup>2</sup> |
| --- | --- | --- | --- | --- |
| Blay et al. 2017 | Soft tissue and visceral sarcomas (STS) | N=12,528<br>Age<sub>median</sub> = 61 (range, 18-101)<br>50.4% Female | Presenting patients to a specialized multidisciplinary tumor board (MDTB) before their initial treatment, with patient outcomes updated in a database. | Presentation to a MDTB before treatment was associated with better compliance to clinical best practices, e.g., rates of biopsy before surgery, adequacy of imaging, quality of initial surgical resection, and fewer reoperations (all p< 0.001). Relapse-free survival was significantly better in patients presented to a MDTB before initiation of treatment, both in univariate and multivariate analysis. |
| Borie et al. 2019 | Interstitial Lung Disease (ILD) | N=95<br>Age<sub>median</sub> = 43 (range, 0-77)<br>44% Female | ILD patients with potential genetic origins are referred by their ILD physician for review by the expert GeneMDD panel who offer diagnostic recommendations and treatment strategies | In addition to diagnostic recommendations, a therapeutic strategy was offered to all (100%) of living patients. Among all 64 patients for whom the geneMDD proposed medication, 63 (93%) eventually received it. |
| Croteau et al. 2016 | Hemophilia | N=60<br>Age<sub>median</sub> = 15 (range, 1-22)<br>3% Female | At a tertiary pediatric center, a quality improvement intervention evaluated and enhanced the transition process for patients with bleeding disorders. The HEMO milestones, offering age-specific steps, were introduced for systematic assessment from infancy to young adulthood during comprehensive clinic visits. | Percentage of patients with documented hemophilia knowledge/skill assessment initially increased from 21% to 100% and was sustained at 97%. Percentage of patients with a documented skill building plan initially increased from 55% to 80% and was further improved to 93%. |
| Han et al. 2016 | Sickle Cell Disease | N = 424<br>Age<sub>median</sub> = 33 (range 25-45)<br>60% Female | In the Sickle Cell Outpatient Center, a new pharmacy service was developed where a clinical pharmacist interacted with patients for ~20 minutes, either before or alongside physicians, addressing drug therapy monitoring, patient education, health counseling, and | The number of pharmacist encounters was associated with 1) improved hydroxyurea dose escalation rate (odds ratio [OR] 1.48, 95% confidence interval [CI] 1.07–2.05, p=0.02); 2) improved immunization completion rates for influenza, PCV13, and PPSV23 (OR 1.38, 95% CI 1.17–1.62, p<0.001) and 3) improved screening for microalbuminuria (OR 2.14, 95% CI 1.60–2.86, p<0.001) and sickle cell retinopathy (OR 1.16, 95% |
|  |  |  | opioid usage evaluations. The pharmacist had the authority to refill non-controlled prescriptions, order vaccines, and made follow-up calls for medication adjustments post-lab reviews. | CI 1.00–1.35, p=0.05). The number of pharmacist encounters was not significantly associated with screening for pulmonary hypertension (p=0.78). |
| Javaud et al. 2018 | Hereditary angioedema | N = 200<br>Age<sub>mean</sub> = 41 (intervention group) and 43 (control group)<br>66% female (intervention group); 60% female (control group) | A dedicated national telephone care-management strategy to reduce resource use during severe angioedema attacks. A national call center was established to provide patients with free, around the clock, phone access to specialist physician advice during a severe attack of hereditary angioedema. | Mean number of hospital admissions for angioedema attacks per patient in the 2-year period was significantly greater in the usual-practice group (mean 0.16) compared to the intervention group (mean 0.03), risk difference: -0.13 (95% confidence interval -0.22 to -0.04; P=.02). For other secondary health outcomes (number of hospital admissions for other causes, ICU admissions per year, number of ED visits, number of intubations, number of interventions by EMS, mortality from angioedema attacks, and number of attacks), there was no statistically significant difference between the intervention and control groups. |
| Kovacikova et al. 2017 | Rare congenital heart defects and other complex cardiac disorders | N = 63<br>Age<sub>median</sub> = 4.3 (range 0-30) | Monthly, multidisciplinary case conferences conducted remotely via videoconferencing. Conferences include presentations of 2-3 local complex cases, consultation from subspecialty experts on diagnostic/therapeutic approach, and review of didactics/care guidelines/literature as needed. | Specialists from the expert center suggested adjustments to the original care plan in 52% of cases. The receiving institution fully accepted changes in 79% of cases, partially accepted changes in 13% of cases, and continued the original plan in 8% of cases. |
| Kreyer et al. 2018 | Ewing sarcoma | N = 481<br>Age<sub>median</sub> = 16.5 (range 0-69.3)<br>39.5% Female | A weekly Interdisciplinary Tumor Board (ITB) that discusses patients upon request from the caring physician or the patient/legal guardian, and reviews patients medical history and imaging to determine treatment recommendations. | A significantly better overall survival was observed in patients with metastatic disease who had been discussed in the ITB and had followed recommendations, compared with patients with metastatic disease who had not been discussed at the ITB (P = 0.028). However, in patients with localized disease, a recommendation from the ITB had no influence on overall survival. |
| Mainous et al. 2018 | Sickle cell disease | N = 71<br>Age<sub>median</sub> = N/A<br>(range 18-45+) | The clinical decision support tool consisted of a best-practice alert that appeared when the EHR was opened and provider education (presentations to faculty and clinic staff about transfusional iron overload.)The alert prompted providers to ask patients if they had a red blood cell transfusion in the past year, and if so to consider ordering a ferritin level. It also displayed the 5 most recent ferritin levels and dates,as well as buttons to Order/Do Not Order a ferritin test | There was a significant increase in the proportion of sickle cell disease patients with an order placed for a ferritin test to assess transfusional iron overload in the intervention group (0% to 44%) and no change in the proportion of patients with a ferritin test ordered in the control group (0% to 0%.) |
| Santoro et al. 2021 | Infantile spasms | N = 17<br>Age<sub>median</sub> = N/A<br>29% Female | An electronic medical record template can be generated by any physician computer user and populated directly into a patient note. The wording generates an assessment and plan in flow sheet format in accordance with the institution's best practices for the evaluation and management of infantile spasms. | Patients who received template-based care had statistically significantly fewer delays in treatment plan (p=0.01), delays in obtaining neurodiagnostic studies (p=0.01), and repeat diagnostic studies needed (p=0.04). There was no statistically significant differences in delays in medication administration (p= 0.10, and delay in discharge (p=0.39) between the two groups. Among 16 resident physicians who used the template, 100% reported the template improved treatment plan comprehension and medication ordering accuracy. |
| Slocum et al. 2018 | Hemophilia | Data regarding the study population was not reported. | The Hemophilia Management Program utilized pharmacists to provide interdisciplinary education, clinical management and continuity of care, and formulary management for hemophilia management. This included developing and distributing educational content to hospital providers and staff, daily assessment of hemophilia patients and attendance at weekly pounds with the hemophilia treatment team, assisting with transitions into and out of inpatient care, and formulary management of factor products. | Consistent reduction in clotting factor utilization, due primarily to reduction of clotting factor dose per patient, resulting in net savings of \$2.7 million for clotting factor expenses without major changes to average length of stay. The program also reduced the need for blood transfusions from 878 to 761 per fiscal year. |
| Turnbull et al. 2021 | Tuberculosis (TB) | N = 424<br>Age<sub>median</sub> = 7.5 and 8 (pre- and post-intervention) (range 0.19 - 16)<br>33% Female | Weekly, virtual MDT discussion between district pediatricians and a tertiary TB team to determine plans for patient diagnosis, management, and follow-up and share this plan with the referring team within 48 hours. | Time to treatment was reduced from a median of 18 days to 13 days after the MDT was established. The proportion of patients who presented with symptoms went from 76% to 55%; the percentage of patients completing treatment with 1 year increased from 94% to 100% and those offered HIV tests increased from 85% to 100%. |
| Wind et al. 2021 | Systemic lupus erythematosus/antiphospholipid syndrome (SLE/APS) | N = 78<br>Age<sub>median</sub>=30-33<br>100% Female | A clinical pathway for evaluation and pre-pregnancy counseling of patients with SLE/APS who desired to become pregnant. Every 2 weeks, all enrolled patients, before and during pregnancy, were discussed in a multidisciplinary team meeting. | Primary outcome of disease-related events (disease flares for patients with SLE and thromboembolic events for patients with APS) was significantly reduced in the pathway cohort compared to a historical cohort (7% vs 28%), mainly determined by a significant reduction in SLE flares. Secondary maternal outcomes (miscarriages, gestational hypertension, severe hypertensive disease, eclampsia, and HELLP syndrome) and fetal outcomes (perinatal death, congenital heart block, preterm birth) were not significantly different. |

<sup>1</sup>All ages reported in years, and all studies assessed gender as male/female only. <sup>2</sup> Additional study outcomes related to diagnosis or clinical care not included in table. Abbreviations: MDTB (multidisciplinary tumor board); Interdisciplinary Tumor Board (ITB)

### Theme 1: Increasing Access to Multidisciplinary Experts

Five studies assessed the impact of multi-disciplinary expert panels to review diagnosis and treatment recommendations.^17–21^ These expert panels, which could be held in-person or virtually, helped experts from multiple disciplines and institutions come together to review patient cases and coordinate patient management plans. These expert panels often recommended therapeutic strategies for patients, which many times involved adjustments to the local care team’s original plans.^17, 18^ These recommendations could increase compliance with clinical best practices^21^, as well as identify patients requiring treatment earlier in the disease process, reduce the time to receive treatment once identified, and increase the number of patients completing treatment.^20^ Expert boards were found to improve relapse-free survival in patients with soft tissue and visceral sarcomas and overall survival in patients with metastatic Ewing’s sarcoma patients, but did not improve overall survival in patients with localized Ewing sarcoma.

Two studies assessed the implementation of clinical pharmacy services in the management of patients. One engaged pharmacists in managing sickle cell disease through in-person services like drug monitoring, patient education, and health counseling.^22^ They handled prescription refills, vaccine orders, and follow-up calls for medication adjustments based on lab results. The program noted improved outcomes in hydroxyurea dose escalation, immunization completion rates, and screenings for microalbuminuria and sickle cell retinopathy due to increased pharmacist interactions. Similarly, another study involved pharmacists in hemophilia management, encompassing interdisciplinary education, clinical oversight, and continuity of care.^23^ Pharmacists developed educational materials, assessed patients daily, and participated in weekly team rounds, aiding in devising drug therapies and patient transitions. The program efficiently reduced clotting factor use and costs by $2.7 million, without major changes to the average length of stay.

One study took a slightly different approach that was more directly targeted towards patient access to a specialist by implementing a new national level hotline. It introduced the SOS-HAE intervention, which is a call center offering 24/7 access to specialized advice for patients.^24^ The call center was staffed by emergency physicians trained in hereditary angioedema management. The ending result was a decrease in hospital admissions for hereditary angioedema attacks. This study offers perspective and lays a foundation for other rare diseases studies in the sense that it suggests that call centers may be effective, depending on the rare diseases, in mitigating patient risk and reducing healthcare costs while still improving patient outcomes.

### Theme 2: Leveraging Technology to Provide Information to Clinicians at Point of Care

Two studies evaluated the use of electronic medical record templates/decision-support tools to improve the quality of care for patients. One introduced an electronic medical record template accessible to physicians for direct inclusion in patient notes.^25^ This template, aligned with institutional best practices, streamlined the evaluation and management of infantile spasms, reducing treatment and diagnostic delays and repetitions. All 16 residents using the template reported full comprehension of the treatment plan and accurate medication ordering. Another study implemented a clinical decision support intervention to enhance adult sickle cell disease care.^26^ The tool, activated upon opening a patient’s electronic health record, prompted providers to inquire about recent red blood cell transfusions and consider ordering a ferritin level test, displaying the latest five ferritin level results. Its introduction led to an increased proportion of ferritin test orders in the intervention group, with no change observed in the control group.

### Theme 3: Standardizing Rare Disease Care

Two studies emphasized a clinical pathway standardizing evaluations to undertake (pregnancy in lupus) or transition process milestones (hemophilia) to achieve certain goals. One involved two patient cohorts, one of which followed a structured pathway involving recommendations such as postponing conception.^27^ The pathway cohort led to a reduction in disease-related events, especially a significant reduction in SLE flares. The reduction in thrombotic events in primary APS pregnancies however was not statistically significant. Overall, this study emphasized that the structured clinical pathway with pre-pregnancy counseling can improve pregnancy outcomes for patients with SLE/APS. Similarly, another study implemented a quality improvement initiative for care transitions for patients with rare bleeding disorders like hemophilia.^28^ By implementing and documenting a longitudinal transition process, specifically the HEMO-Milestones tool, patient knowledge and skills related to hemophilia could be assessed. Overall, both studies introduced a pathway to assess patient transition and involved multidisciplinary teams. Both pathways also consider age-appropriate milestones, the first study emphasizes infancy to young adulthood and the second study outlines goals for both pediatric and adult patients. Finally, both studies consider the significance of documenting patient progress and measuring milestone achievement.

## Discussion

The objective of this scoping review was to identify health services interventions that were aimed at increasing rare disease patients’ quality of care by improving care coordination, care transitions, and adherence to care guidelines and best practices. Despite the high unmet need to improve healthcare delivery for individuals with rare disease, only 12 full-text, peer reviewed studies with published results were identified using our search terms focused on rare diseases generally. However, we were still able to identify common intervention techniques across studies with high potential to improve care across rare diseases more broadly.

Across interventions, we recognized three main trends that contributed to more efficient healthcare delivery. First, research frequently aims to improve patient access to specialists and to involve team members with the necessary knowledge to elevate the quality of care. This addresses a primary challenge in rare disease treatment: the scarcity of specialists equipped with the training and understanding required for these complex conditions.^29^ Telehealth solutions and call centers, which are increasingly trialed in interventional studies, tend to reduce geographical barriers and quickly increase access to medical expertise. Research concluded in several countries have pointed towards a need for more training for health professionals in order for information to be communicated effectively.^30^

The second trend in retrieved articles was providing point-of-care or attending physicians with integrated technology, such as electronic medical records, was vital for advancing the care they delivered. The second trend addresses the critical need for the integration and utilization of technology, which is essential for enhancing diagnostic accuracy and treatment efficacy in the realm of rare diseases.^31^ Now more than ever, with the boom in AI, research points towards a path to speed up rare disease diagnoses as well as create decision support systems. The advancement of technology has and will continue to make a substantial positive impact on the lives of patients with rare diseases when implemented in an ethical manner. Patients with rare diseases deserve not only quality care, but also rapid care– technology can speed up the process of creating personalized treatment plans after expert consultations.^32^

The third trend across studies was the need to standardize care practices to minimize variations in treatment quality, ensuring all care meets the highest standards. This trend underscores the significance of establishing standardized care protocols, particularly vital in a field where consistency, structured approaches, and comprehensive disease knowledge are often lacking.^33^ The highlighted intervention trends emphasize the challenges patients with rare diseases face in accessing specialized expertise and receiving appropriate care. Moreover, the studies emphasized that the quality of life for children and adults with rare diseases must also be improved through standardization of care. By reducing variation across rare diseases, patients will consistently receive evidence-based treatments. This will require boosting coordination of care among healthcare providers. Overall, standardization ensures that the correct treatments are given based on recommendations from experts in their respective fields.^34^

The findings of common themes underlying multiple, successful rare disease health services interventions identified in this scoping review suggest a high likelihood of the transferability and scalability of these intervention approaches. However, our study does have several limitations. The primary limitation is that we employed “rare disease” as a key term for feasibility purposes rather than compiling a list of individual rare diseases. This may have resulted in the omission of some studies that did not explicitly mention that they were addressing a “rare disease” or relevant synonym. However, since our review aim was to

identify interventions that could be translated across rare diseases, we expect the most relevant papers would include these terms. Further limitations on the generalizability of our findings arises from the fact that most of the included studies were conducted in Western, high-income countries. Our inclusion of only English-language papers could have unintentionally excluded papers from other countries, including lower resource settings that may require other intervention methodologies. Lastly, another potential limitation is the presence of publication bias. It is likely that interventions with negative or inconclusive outcomes may not be published, thereby skewing the representation of effective interventions in our review. This bias could limit our understanding of the full spectrum of interventions and their outcomes for rare diseases.

Despite these limitations, this was a rigorous review of health services interventions for rare diseases that identified several efficacious interventions that could be spread to other rare disease contexts. This review, while likely not exhaustive of every rare disease health services intervention, still underscores the significance and need for more research focused on addressing common barriers to rare disease care. Collaborative research to improve healthcare delivery systems is imperative to increase rare disease patients’ access to high-quality care and new therapeutic advances. While all studies identified in this scoping review studied a single rare disease or a group of closely related diseases, designing studies that encompass individuals with a variety of rare diseases may offer a more effective strategy for identifying broadly applicable solutions across diverse patient demographics.^35^ Adopting this cross-disorder approach to healthcare delivery innovation could enable transformative advancements in the care and treatment of the rare disease community as a whole.

## Data Availability

As a scoping review, no new data was generated. A full list of retrieved studies assessed for inclusion/exclusion in the review is available upon reasonable request to the authors.

## Declarations

### Ethics approval and consent to participate

As a scoping literature review without study participants, no ethics approval or consent was required.

### Consent for publication

N/A.

### Author Contributions

Conceptualization: LVI, SA, VM. Data curation: ML. Methodology: ML, VM. Supervision: VM. Investigation: CC, SW, LVI, AT, SA, VM. Formal Analysis, Visualization, and Writing – original draft: CC, SW, VM. Writing – review and editing: LVI, AT, SA, ML.

## Acknowledgements

N/A

## Availability of data and materials

Not applicable.

## Appendix A. Search Strategy

**Supplemental Table 1.**
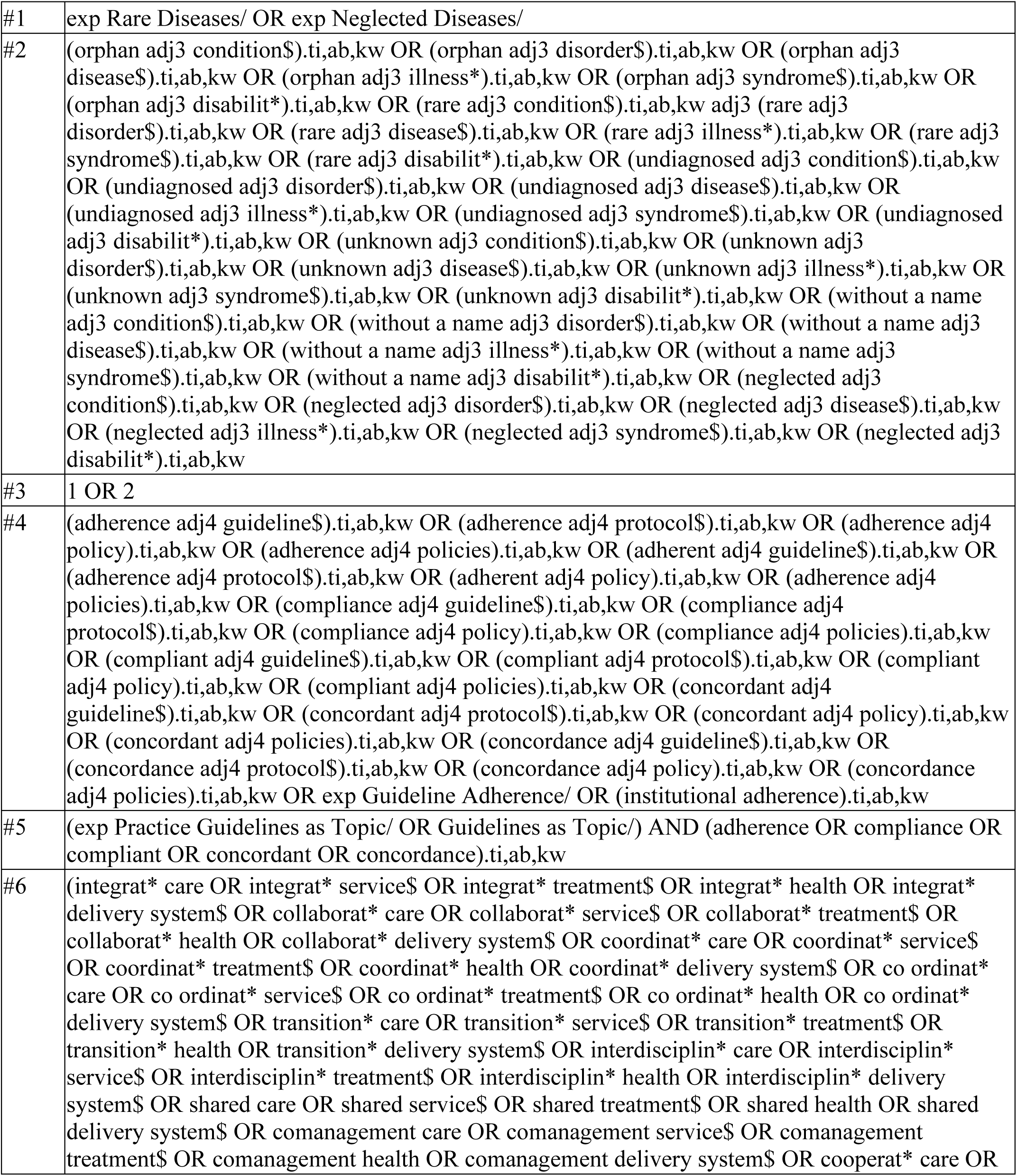
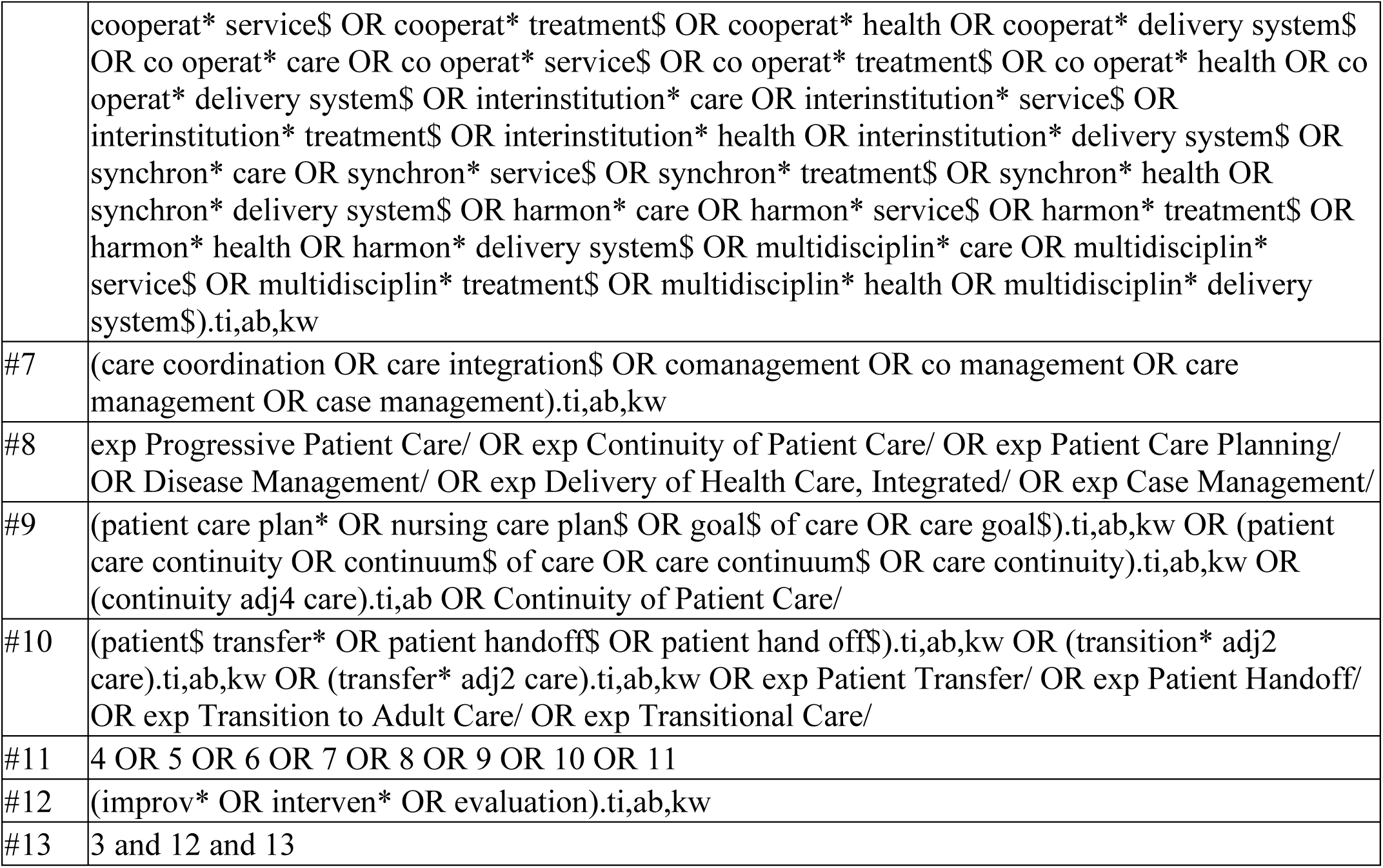
Full Search Strategy for MEDLINE Searched on 4/20/22.

**Supplemental Table 2.** Full Search Strategy for Embase.com Searched on 4/20/22.

|  |  |
| --- | --- |
| #1 | 'Rare Disease'/exp OR 'Neglected Disease'/exp OR 'undiagnosed disease'/exp |
| #2 | (orphan NEAR/3 condition\$):ti,ab,kw OR (orphan NEAR/3 disorder\$):ti,ab,kw OR (orphan NEAR/3 disease\$):ti,ab,kw OR (orphan NEAR/3 illness*):ti,ab,kw OR (orphan NEAR/3 syndrome\$):ti,ab,kw OR (orphan NEAR/3 disabilit*):ti,ab,kw OR (rare NEAR/3 condition\$):ti,ab,kw OR (rare NEAR/3 disorder\$):ti,ab,kw OR (rare NEAR/3 disease\$):ti,ab,kw OR (rare NEAR/3 illness*):ti,ab,kw OR (rare NEAR/3 syndrome\$):ti,ab,kw OR (rare NEAR/3 disabilit*):ti,ab,kw OR (undiagnosed NEAR/3 condition\$):ti,ab,kw OR (undiagnosed NEAR/3 disorder\$):ti,ab,kw OR (undiagnosed NEAR/3 disease\$):ti,ab,kw OR (undiagnosed NEAR/3 illness*):ti,ab,kw OR (undiagnosed NEAR/3 syndrome\$):ti,ab,kw OR (undiagnosed NEAR/3 disabilit*):ti,ab,kw OR (unknown NEAR/3 condition\$):ti,ab,kw OR (unknown NEAR/3 disorder\$):ti,ab,kw OR (unknown NEAR/3 disease\$):ti,ab,kw OR (unknown NEAR/3 illness*):ti,ab,kw OR (unknown NEAR/3 syndrome\$):ti,ab,kw OR (unknown NEAR/3 disabilit*):ti,ab,kw OR (neglected NEAR/3 condition\$):ti,ab,kw OR (neglected NEAR/3 disorder\$):ti,ab,kw OR (neglected NEAR/3 disease\$):ti,ab,kw OR (neglected NEAR/3 illness*):ti,ab,kw OR (neglected NEAR/3 syndrome\$):ti,ab,kw OR (neglected NEAR/3 disabilit*):ti,ab,kw OR ('without a name':ti,ab,kw AND (condition\$ OR disorder\$ OR disease\$ OR illness* OR syndrome\$ OR disabilit*):ab,ti,kw) |
| #3 | #1 OR #2 |
| #4 | (adherence NEAR/4 guideline\$):ti,ab,kw OR (adherence NEAR/4 protocol\$):ti,ab,kw OR (adherence NEAR/4 policy):ti,ab,kw OR (adherence NEAR/4 policies):ti,ab,kw OR (adherent NEAR/4 guideline\$):ti,ab,kw OR (adherence NEAR/4 protocol\$):ti,ab,kw OR (adherent NEAR/4 policy):ti,ab,kw OR (adherence NEAR/4 policies):ti,ab,kw OR (compliance NEAR/4 guideline\$):ti,ab,kw OR (compliance NEAR/4 protocol\$):ti,ab,kw OR (compliance NEAR/4 policy):ti,ab,kw OR (compliance NEAR/4 policies):ti,ab,kw OR (compliant NEAR/4 guideline\$):ti,ab,kw OR (compliant NEAR/4 protocol\$):ti,ab,kw OR (compliant NEAR/4 policy):ti,ab,kw OR (compliant NEAR/4 policies):ti,ab,kw OR (concordant NEAR/4 guideline\$):ti,ab,kw OR (concordant NEAR/4 protocol\$):ti,ab,kw OR (concordant NEAR/4 policy):ti,ab,kw OR (concordant NEAR/4 policies):ti,ab,kw OR (concordance NEAR/4 guideline\$):ti,ab,kw OR (concordance NEAR/4 protocol\$):ti,ab,kw OR (concordance NEAR/4 policy):ti,ab,kw OR (concordance NEAR/4 policies):ti,ab,kw OR 'protocol compliance'/exp OR 'institutional adherence':ab,ti,kw |
| #5 | ('guideline directed medical therapy'/exp OR 'practice guideline'/de OR 'clinical protocol'/de OR 'nursing protocol'/exp) AND (adherence OR compliance OR compliant OR concordant OR concordance):ab,ti,kw |
| #6 | ('integrat* care' OR 'integrat* service\$' OR 'integrat* treatment\$' OR 'integrat* health' OR 'integrat* delivery system\$' OR 'collaborat* care' OR 'collaborat* service\$' OR 'collaborat* treatment\$' OR 'collaborat* health' OR 'collaborat* delivery system\$' OR 'coordinat* care' OR 'coordinat* service\$' OR 'coordinat* treatment\$' OR 'coordinat* health' OR 'coordinat* delivery system\$' OR 'co ordinat* care' OR 'co ordinat* service\$' OR 'co ordinat* treatment\$' OR 'co ordinat* health' OR 'co ordinat* delivery system\$' OR 'transition* care' OR 'transition* service\$' OR 'transition* treatment\$' OR 'transition* health' OR 'transition* delivery system\$' OR 'interdisciplin* care' OR 'interdisciplin* service\$' OR 'interdisciplin* treatment\$' OR 'interdisciplin* health' OR 'interdisciplin* delivery system\$' OR 'shared care' OR 'shared service\$' OR 'shared treatment\$' OR 'shared health' OR 'shared delivery system\$' OR 'comanagement care' OR 'comanagement service\$' OR 'comanagement treatment\$' OR 'comanagement health' OR 'comanagement delivery system\$' OR 'cooperat* care' OR 'cooperat* service\$' OR 'cooperat* treatment\$' OR 'cooperat* health' OR 'cooperat* delivery system\$' OR 'co operat* care' OR 'co operat* service\$' OR 'co operat* treatment\$' OR 'co operat* health' OR 'co operat* delivery system\$' OR 'interinstitution* care' OR 'interinstitution* service\$' OR 'interinstitution* treatment\$' OR 'interinstitution* health' OR 'interinstitution* delivery system\$' OR 'synchron* care' OR 'synchron* service\$' OR 'synchron* treatment\$' OR 'synchron* health' OR 'synchron* delivery system\$' OR 'harmon* care' OR 'harmon* service\$' OR 'harmon* treatment\$' OR 'harmon* health' OR 'harmon* delivery system\$' OR 'multidisciplin* care' OR 'multidisciplin* service\$' OR 'multidisciplin* treatment\$' OR 'multidisciplin* health' OR 'multidisciplin* delivery system\$'):ab,ti,kw OR 'collaborative care'/exp OR 'collaborative care team'/exp OR 'collaborative care model'/exp OR 'coordinated care'/exp OR 'interdisciplinary care'/exp OR 'interdisciplinary team'/exp OR 'shared care'/exp OR 'multidisciplinary team'/exp OR 'multidisciplinary care'/exp OR 'multidisciplinary approach'/exp OR 'multidisciplinary management'/exp |
| #7 | ('care coordination' OR 'care integration\$' OR comanagement OR 'co management' OR 'care management' OR 'case management'):ab,ti,kw |
| #8 | 'Progressive Patient Care'/exp OR 'Patient Care'/de OR 'Patient Care Planning'/exp OR 'Disease Management'/de OR 'Integrated Care'/exp OR 'integrated health care system'/exp OR 'integrated care pathway'/exp OR 'care coordination'/exp OR 'case management'/exp OR 'disease management program'/exp |
| #9 | ('patient care plan*' OR 'nursing care plan\$' OR 'goal\$ of care' OR 'care goal\$' OR 'patient care continuity' OR 'continuum\$ of care' OR 'care continuum\$' OR 'care continuity'):ab,ti,kw OR (continuity NEAR/4 care):ab,ti OR 'nursing care plan'/exp OR 'patient care planning'/exp |
| #10 | ('patient\$ transfer*' OR 'patient handoff\$' OR 'patient hand off\$'):ab,ti,kw OR (transition* NEAR/2 care):ab,ti,kw OR (transfer* NEAR/2 care):ab,ti,kw OR 'clinical handover'/de OR 'transition to adult care'/exp OR 'transitional care'/exp |
| #11 | #4 OR #5 OR #6 OR #7 OR #8 OR #9 OR #10 OR #11 |
| #12 | (improv* OR interven* OR evaluation):ab,ti,kw OR 'intervention'/exp |
| #13 | #3 AND #11 AND #12 |

**Supplemental Table 3.**
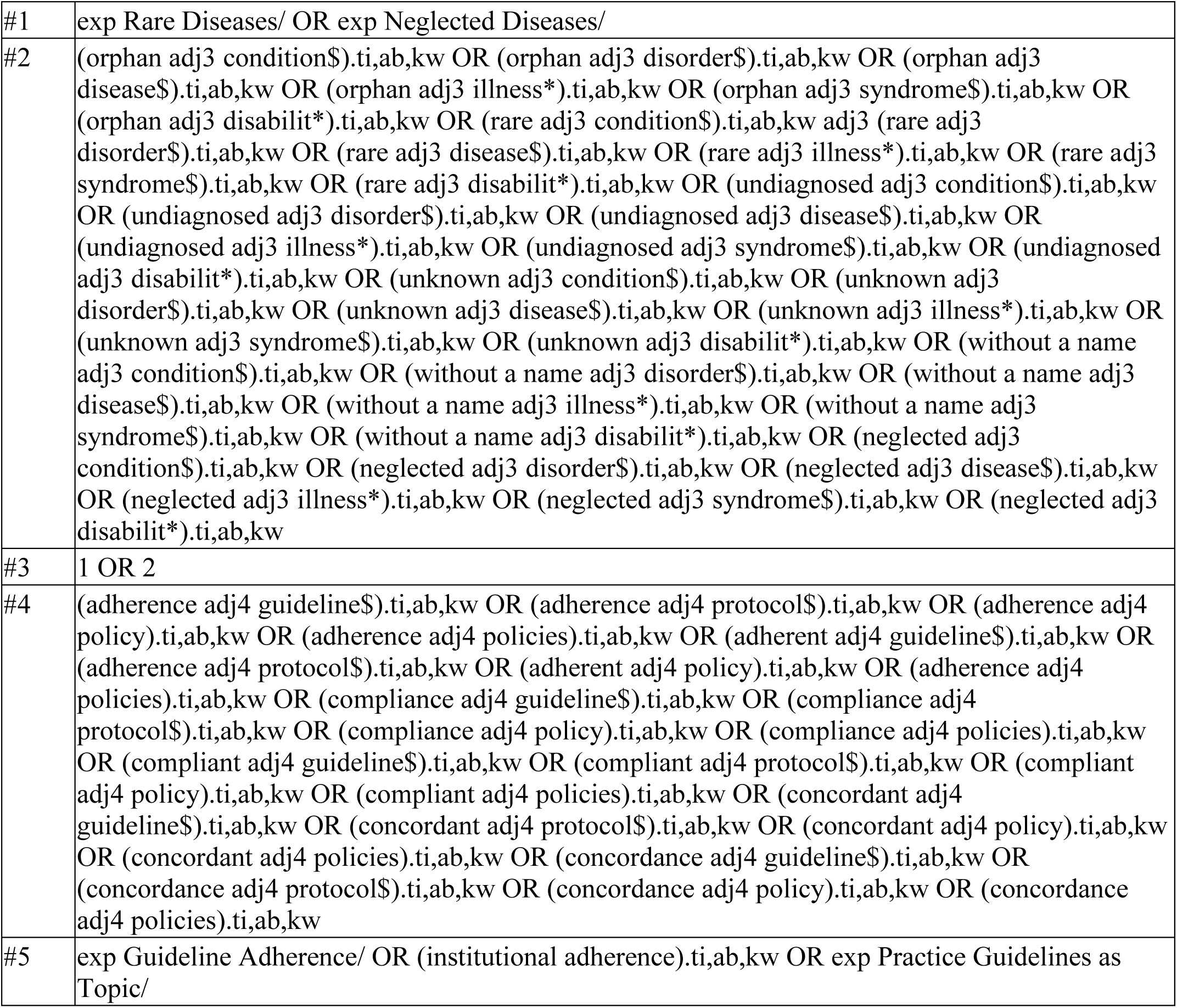
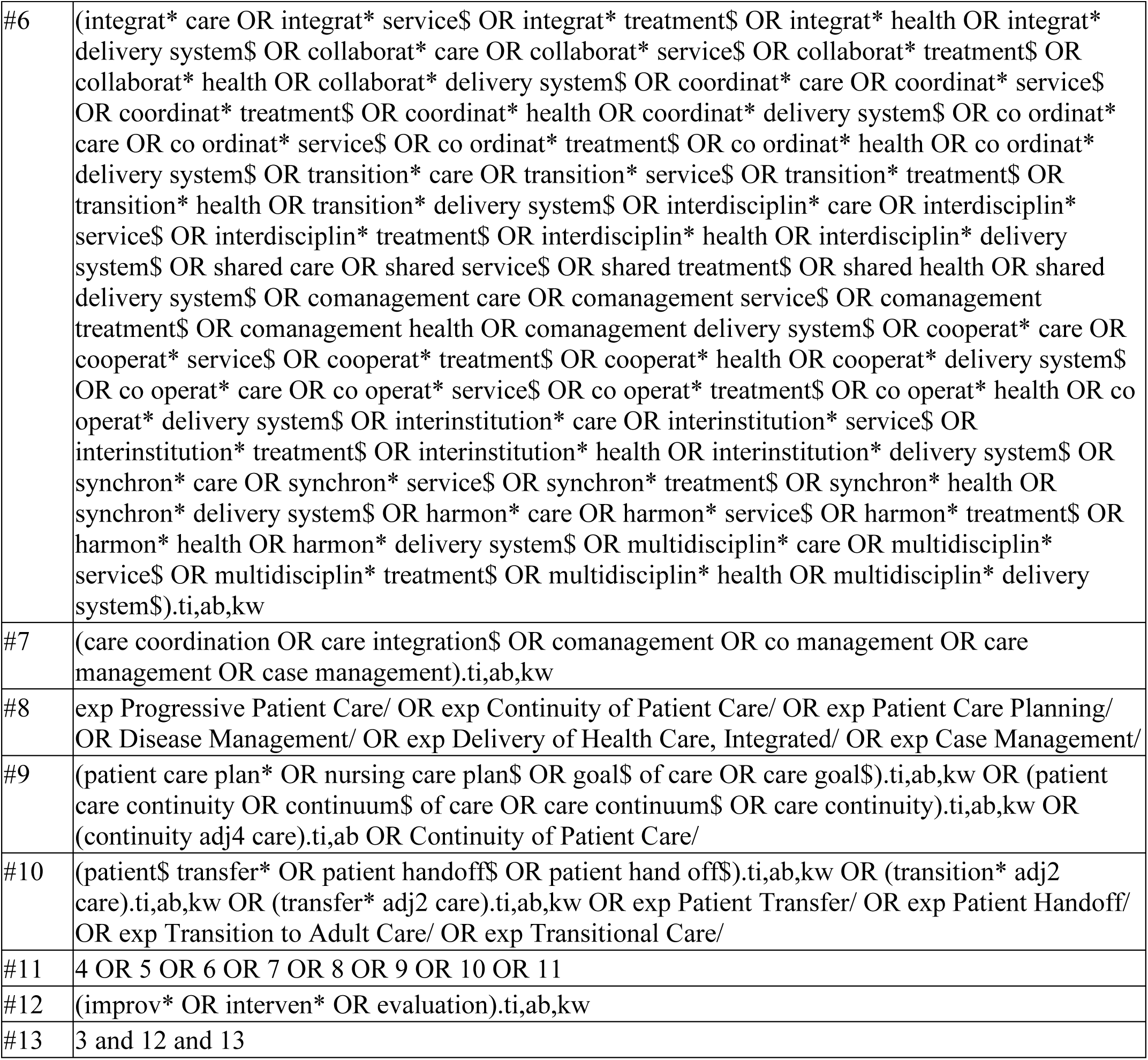
Full Search Strategy for Cochrane Searched on 4/20/22.

**Supplemental Table 4.** Full Search Strategy for Web of Science Searched on 4/19/22.

|  |  |
| --- | --- |
| #1 | TS=((orphan NEAR/3 condition\$) OR (orphan NEAR/3 disorder\$) OR (orphan NEAR/3 disease\$) OR (orphan NEAR/3 illness*) OR (orphan NEAR/3 syndrome\$) OR (orphan NEAR/3 disabilit*) OR (rare NEAR/3 condition\$) NEAR/3 (rare NEAR/3 disorder\$) OR (rare NEAR/3 disease\$) OR (rare NEAR/3 illness*) OR (rare NEAR/3 syndrome\$) OR (rare NEAR/3 disabilit*) OR (undiagnosed NEAR/3 condition\$) OR (undiagnosed NEAR/3 disorder\$) OR (undiagnosed NEAR/3 disease\$) OR (undiagnosed NEAR/3 illness*) OR (undiagnosed NEAR/3 syndrome\$) OR (undiagnosed NEAR/3 disabilit*) OR (unknown NEAR/3 condition\$) OR (unknown NEAR/3 disorder\$) OR (unknown NEAR/3 disease\$) OR (unknown NEAR/3 illness*) OR (unknown NEAR/3 syndrome\$) OR (unknown NEAR/3 disabilit*) OR (without a name NEAR/3 condition\$) |
| | OR (without a name NEAR/3 disorder\$) OR (without a name NEAR/3 disease\$) OR (without a name NEAR/3 illness*) OR (without a name NEAR/3 syndrome\$) OR (without a name NEAR/3 disabilit*) OR (neglected NEAR/3 condition\$) OR (neglected NEAR/3 disorder\$) OR (neglected NEAR/3 disease\$) OR (neglected NEAR/3 illness*) OR (neglected NEAR/3 syndrome\$) OR (neglected NEAR/3 disabilit*) |
| #2 | TS=((adherence NEAR/4 guideline\$) OR (adherence NEAR/4 protocol\$) OR (adherence NEAR/4 policy) OR (adherence NEAR/4 policies) OR (adherent NEAR/4 guideline\$) OR (adherence NEAR/4 protocol\$) OR (adherent NEAR/4 policy) OR (adherence NEAR/4 policies) OR (compliance NEAR/4 guideline\$) OR (compliance NEAR/4 protocol\$) OR (compliance NEAR/4 policy) OR (compliance NEAR/4 policies) OR (compliant NEAR/4 guideline\$) OR (compliant NEAR/4 protocol\$) OR (compliant NEAR/4 policy) OR (compliant NEAR/4 policies) OR (concordant NEAR/4 guideline\$) OR (concordant NEAR/4 protocol\$) OR (concordant NEAR/4 policy) OR (concordant NEAR/4 policies) OR (concordance NEAR/4 guideline\$) OR (concordance NEAR/4 protocol\$) OR (concordance NEAR/4 policy) OR (concordance NEAR/4 policies)) |
| #3 | TS=("guideline adherence" OR "institutional adherence") |
| #4 | TS=("integrat* care" OR "integrat* service\$" OR "integrat* treatment\$" OR "integrat* health" OR "integrat* delivery system\$" OR "collaborat* care" OR "collaborat* service\$" OR "collaborat* treatment\$" OR "collaborat* health" OR "collaborat* delivery system\$" OR "coordinat* care" OR "coordinat* service\$" OR "coordinat* treatment\$" OR "coordinat* health" OR "coordinat* delivery system\$" OR "co ordinat* care" OR "co ordinat* service\$" OR "co ordinat* treatment\$" OR "co ordinat* health" OR "co ordinat* delivery system\$" OR "transition* care" OR "transition* service\$" OR "transition* treatment\$" OR "transition* health" OR "transition* delivery system\$" OR "interdisciplin* care" OR "interdisciplin* service\$" OR "interdisciplin* treatment\$" OR "interdisciplin* health" OR "interdisciplin* delivery system\$" OR "shared care" OR "shared service\$" OR "shared treatment\$" OR "shared health" OR "shared delivery system\$" OR "comanagement care" OR "comanagement service\$" OR "comanagement treatment\$" OR "comanagement health" OR "comanagement delivery system\$" OR "cooperat* care" OR "cooperat* service\$" OR "cooperat* treatment\$" OR "cooperat* health" OR "cooperat* delivery system\$" OR "co operat* care" OR "co operat* service\$" OR "co operat* treatment\$" OR "co operat* health" OR "co operat* delivery system\$" OR "interinstitution* care" OR "interinstitution* service\$" OR "interinstitution* treatment\$" OR "interinstitution* health" OR "interinstitution* delivery system\$" OR "synchron* care" OR "synchron* service\$" OR "synchron* treatment\$" OR "synchron* health" OR "synchron* delivery system\$" OR "harmon* care" OR "harmon* service\$" OR "harmon* treatment\$" OR "harmon* health" OR "harmon* delivery system\$" OR "multidisciplin* care" OR "multidisciplin* service\$" OR "multidisciplin* treatment\$" OR "multidisciplin* health" OR "multidisciplin* delivery system\$") |
| #5 | TS=("care coordination" OR "care integration\$" OR comanagement OR co management OR "care management" OR "case management" OR "disease management" OR "progressive patient care") |
| #6 | TS=("patient care plan*" OR "nursing care plan\$" OR "goal\$ of care" OR "care goal\$" OR "continuum\$ of care" OR "care continuum\$" OR "care continuity" OR (continuity NEAR/4 care)) |
| #7 | TS=("patient\$ transfer*" OR "patient handoff\$" OR "patient hand off\$" OR (transition* NEAR/2 care) OR (transfer* NEAR/2 care)) |
| #8 | #2 OR #3 OR #4 OR #5 OR #6 OR #7 |
| #9 | TS=(improv* OR interven* OR evaluation) |
| #10 | #1 AND #8 AND #9 |

**Supplemental Table 5.**
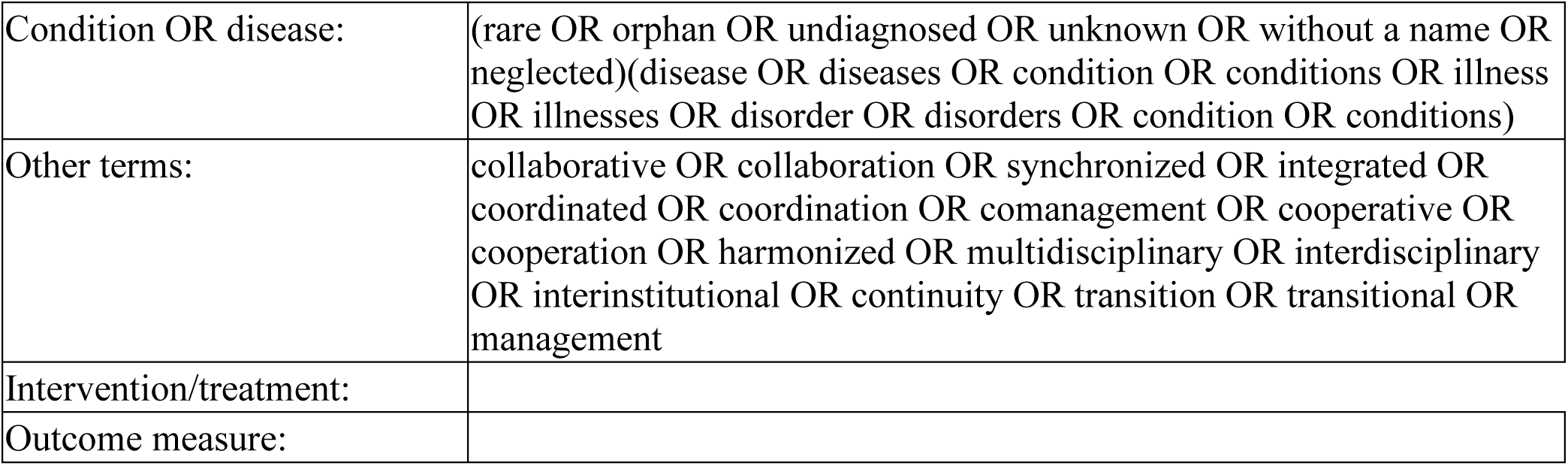
Full Search Strategy for ClinicalTrials.gov Searched on 4/19/22.

## References

1. RDD-FAQ-2019.pdf [Internet]. [cited 2024 Jan 29]. Available from: https://rarediseases.org/wp-content/uploads/2019/01/RDD-FAQ-2019.pdf

2. Reinhard C, Bachoud-Lévi AC, Bäumer T, Bertini E, Brunelle A, Buizer AI, et al. The European Reference Network for Rare Neurological Diseases. Front Neurol [Internet]. 2021 [cited 2024 Jan 29];11. Available from: https://www.frontiersin.org/articles/10.3389/fneur.2020.616569

3. Cortese S, Song M, Farhat LC, Yon DK, Lee SW, Kim MS, et al. Incidence, prevalence, and global burden of ADHD from 1990 to 2019 across 204 countries: data, with critical re-analysis, from the Global Burden of Disease study. Mol Psychiatry. 2023 Sep 8;1–8.

4. Slade A, Isa F, Kyte D, Pankhurst T, Kerecuk L, Ferguson J, et al. Patient reported outcome measures in rare diseases: a narrative review. Orphanet J Rare Dis. 2018 Apr 23;13(1):61.

5. Bryson B, Bogart K, Atwood M, Fraser K, Locke T, Pugh K, et al. Navigating the unknown: A content analysis of the unique challenges faced by adults with rare diseases. J Health Psychol. 2021 Apr;26(5):623–35.

6. Munro M, Cook AM, Bogart KR. An inductive qualitative content analysis of stigma experienced by people with rare diseases. Psychol Health. 2022 Aug;37(8):948–63.

7. Closing the Quality Gap: A Critical Analysis of Quality Improvement Strategies, Volume 7-- Care Coordination.

8. Walton H, Simpson A, Ramsay AIG, Hunter A, Jones J, Ng PL, et al. Development of models of care coordination for rare conditions: a qualitative study. Orphanet J Rare Dis. 2022 Feb 14;17(1):49.

9. Bogart K, Hemmesch A, Barnes E, Blissenbach T, Beisang A, Engel P, et al. Healthcare access, satisfaction, and health-related quality of life among children and adults with rare diseases. Orphanet J Rare Dis. 2022 May 12;17(1):196.

10. von der Lippe C, Diesen PS, Feragen KB. Living with a rare disorder: a systematic review of the qualitative literature. Mol Genet Genomic Med. 2017 Jul 23;5(6):758–73.

11. Stepien KM, Kieć-Wilk B, Lampe C, Tangeraas T, Cefalo G, Belmatoug N, et al. Challenges in Transition From Childhood to Adulthood Care in Rare Metabolic Diseases: Results From the First Multi-Center European Survey. Front Med [Internet]. 2021 [cited 2024 Jan 30];8. Available from: https://www.frontiersin.org/articles/10.3389/fmed.2021.652358

12. Sandquist M, Davenport T, Monaco J, Lyon ME. The Transition to Adulthood for Youth Living with Rare Diseases. Children. 2022 May 12;9(5):710.

13. Valdez R. Public Health and Rare Diseases: Oxymoron No More. Prev Chronic Dis [Internet]. 2016 [cited 2024 Jan 30];13. Available from: https://www.cdc.gov/pcd/issues/2016/15_0491.htm

14. Van Groenendael S, Giacovazzi L, Davison F, Holtkemper O, Huang Z, Wang Q, et al. High quality, patient centred and coordinated care for Alstrom syndrome: a model of care for an ultra-rare disease. Orphanet J Rare Dis. 2015 Nov 24;10(1):149.

15. Arksey H, O’Malley L. Scoping Studies: Towards a Methodological Framework. Int J Soc Res Methodol Theory Pract. 2005;8(1):19–32.

16. Covidence - Better systematic review management [Internet]. [cited 2024 Jan 30]. Available from: https://www.covidence.org/

17. Borie R, Kannengiesser C, Gouya L, Dupin C, Amselem S, Ba I, et al. Pilot experience of multidisciplinary team discussion dedicated to inherited pulmonary fibrosis. Orphanet J Rare Dis. 2019 Dec;14(1):280.

18. Kovacikova L, Zahorec M, Skrak P, Hanna BD, Lee Vogel R. Transatlantic medical consultation and second opinion in pediatric cardiology has benefit past patient care: A case study in videoconferencing: KOVACIKOVA et al. Congenit Heart Dis. 2017 Jul;12(4):491–6.

19. Kreyer J, Ranft A, Timmermann B, Juergens H, Jung S, Wiebe K, et al. Impact of the Interdisciplinary Tumor Board of the Cooperative Ewing Sarcoma Study Group on local therapy and overall survival of Ewing sarcoma patients after induction therapy. Pediatr Blood Cancer. 2018 Dec;65(12):e27384.

20. Turnbull L, Bell C, Davies S, Child F. Delivering tertiary tuberculosis care virtually. Arch Dis Child. 2021 Dec;106(12):1226–8.

21. Blay JY, Soibinet P, Penel N, Bompas E, Duffaud F, Stoeckle E, et al. Improved survival using specialized multidisciplinary board in sarcoma patients. Ann Oncol. 2017 Nov;28(11):2852–9.

22. Han J, Bhat S, Gowhari M, Gordeuk VR, Saraf SL. Impact of a Clinical Pharmacy Service on the Management of Patients in a Sickle Cell Disease Outpatient Center. Pharmacother J Hum Pharmacol Drug Ther. 2016 Nov;36(11):1166–72.

23. Slocum GW, Peksa GD, Webb TA, Lat I. Implementation of a hemophilia management program improves clinical outcomes. JACCP J Am Coll Clin Pharm. 2019 Jun;2(3):236–42.

24. Javaud N, Fain O, Durand-Zaleski I, Launay D, Bouillet L, Gompel A, et al. Specialist Advice Support for Management of Severe Hereditary Angioedema Attacks: A Multicenter Cluster-Randomized Controlled Trial. Ann Emerg Med. 2018 Aug;72(2):194–203.e1.

25. Santoro JD, Sandoval Karamian AG, Ruzhnikov M, Brimble E, Chadwick W, Wusthoff CJ. Use of electronic medical record templates improves quality of care for patients with infantile spasms. Health Inf Manag J. 2021 Jan;50(1–2):47–54.

26. Mainous AG, Carek PJ, Lynch K, Tanner RJ, Hulihan MM, Baskin J, et al. Effectiveness of Clinical Decision Support Based Intervention in the Improvement of Care for Adult Sickle Cell Disease Patients in Primary Care. J Am Board Fam Med. 2018 Sep;31(5):812–6.

27. Wind M, Hendriks M, Van Brussel BTJ, Eikenboom J, Allaart CF, Lamb HJ, et al. Effectiveness of a multidisciplinary clinical pathway for women with systemic lupus erythematosus and/or antiphospholipid syndrome. Lupus Sci Med. 2021 May;8(1):e000472.

28. Croteau SE, Padula M, Quint K, D’Angelo L, Neufeld EJ. Center-Based Quality Initiative Targets Youth Preparedness for Medical Independence: *HEMO-Milestones Tool* in a Comprehensive Hemophilia Clinic Setting: Quality Initiative Targets Youth Transition Process. Pediatr Blood Cancer. 2016 Mar;63(3):499–503.

29. Elliott E, Zurynski Y. Rare diseases are a “common” problem for clinicians. Aust Fam Physician. 2015 Sep;44(9):630–3.

30. Merker VL, Dai A, Radtke HB, Knight P, Jordan JT, Plotkin SR. Increasing access to specialty care for rare diseases: a case study using a foundation sponsored clinic network for patients with neurofibromatosis 1, neurofibromatosis 2, and schwannomatosis. BMC Health Serv Res. 2018 Aug 29;18:668.

31. Ronicke S, Hirsch MC, Türk E, Larionov K, Tientcheu D, Wagner AD. Can a decision support system accelerate rare disease diagnosis? Evaluating the potential impact of Ada DX in a retrospective study. Orphanet J Rare Dis. 2019 Mar 21;14(1):69.

32. Wojtara M, Rana E, Rahman T, Khanna P, Singh H. Artificial intelligence in rare disease diagnosis and treatment. Clin Transl Sci. 2023 Nov;16(11):2106–11.

33. Adachi T, El-Hattab AW, Jain R, Nogales Crespo KA, Quirland Lazo CI, Scarpa M, et al. Enhancing Equitable Access to Rare Disease Diagnosis and Treatment around the World: A Review of Evidence, Policies, and Challenges. Int J Environ Res Public Health. 2023 Mar 8;20(6):4732.

34. Pai M, Yeung CHT, Akl EA, Darzi A, Hillis C, Legault K, et al. Strategies for eliciting and synthesizing evidence for guidelines in rare diseases. BMC Med Res Methodol. 2019 Mar 28;19:67.

35. Depping MK, Uhlenbusch N, Härter M, Schramm C, Löwe B. Efficacy of a Brief, Peer-Delivered Self-management Intervention for Patients With Rare Chronic Diseases: A Randomized Clinical Trial. JAMA Psychiatry. 2021 Jun 1;78(6):607–15.

